# Portable and label-free quantitative Loop-mediated Isothermal Amplification (qLAMP) for reliable COVID-19 diagnostics in 3 minutes: An Arduino-based detection system assisted by a pH microelectrode

**DOI:** 10.1101/2021.05.23.21256350

**Authors:** Mario Moisés Alvarez, Sergio Bravo-González, Everardo González-González, Grissel Trujillo-de Santiago

## Abstract

Loop-mediated isothermal amplification (LAMP) has been recently studied as an alternative method for cost-effective diagnostics in the context of the current COVID-19 pandemic. Recent reports document that LAMP-based diagnostic methods have a comparable sensitivity and specificity to that of RT-qPCR. We report the use of a portable Arduino-based LAMP-based amplification system assisted by pH microelectrodes for the accurate and reliable diagnosis of SARS-CoV-2 during the first 3 minutes of the amplification reaction. We show that this simple system enables a straightforward discrimination between samples containing or not containing artificial SARS-CoV-2 genetic material in the range of 10 to 10,000 copies per 50 µL of reaction mix. We also spiked saliva samples with SARS-CoV-2 synthetic material and corroborated that the LAMP reaction can be successfully monitored in real time using microelectrodes in saliva samples as well. These results may have profound implications for the design of real-time and portable quantitative systems for the reliable detection of viral pathogens including SARS-CoV-2.

## Introduction

At present, nearly 150 million people have been diagnosed with COVID-19 (+) worldwide, and the disease has killed more than 3 million people [1], making it the most lethal infectious disease in a century. COVID-19 has laid bare the severe limitations in our capacities to respond to epidemic emergencies and has made clear that we must expand and strengthen the portfolio of available tools for effective diagnostics of infectious diseases [2] both now and in the future.

The retro-transcriptase quantitative polymerase chain reaction (RT-qPCR) is currently the gold standard methodology for COVID-19 detection [2–5]. Despite its unquestionable accuracy and robustness, qPCR-based methods have some serious limitations (e.g., lack of portability, dependence on centralized facilities, need for technical expertise to conduct RT-qPCR testing, and high infrastructural and operative costs) that prevent the provision of cost-effective and massive diagnostics during this time of COVID-19 [2,6,7].

One potential alternative to RT-qPCR could be Loop-mediated Isothermal Amplification (LAMP), which has been extensively studied in recent reports as a cost-effective diagnostic system in the context of the current COVID-19 pandemic [8–16]. Overall, these reports have documented that LAMP-based methods have comparable sensitivity and specificity to that of RT-qPCR. More importantly, LAMP methods provide attractive advantages over RT-qPCR methods in terms of lower capital and operating costs. LAMP is an isothermal method, so it frees the user from the need for a costly thermal cycler. In addition, LAMP methods that exhibit excellent sensibility and acceptable specificity have been described for the extraction-free analysis of nasopharyngeal and even saliva samples [15,17,18]. The release from the need for an extraction stage further reduces the cost of LAMP methodology.

Most LAMP-based methods rely on a discrimination between positive and negative samples due to a change in the pH associated with an acidification of the reaction mix due to the liberation of hydrogen ions (H_3_O^+^) inherent in the amplification [15,17,19]. In a weakly buffered reaction medium, the change in pH may translate into a change in color if a pH indicator is added to the reaction mix. A frequent embodiment of colorimetric LAMP uses phenol red as the pH indicator [17,19–21] and a weak buffer that contains mainly Tris-HCl, (NH4)_2_SO_4_ (ammonium sulfate), and KCl (potassium chloride).

However, colorimetric LAMP methods also have some limitations. One is that the adequate performance of colorimetric LAMP mediated by phenol red depends on the initial pH of the sample. This is particularly relevant in extraction-free applications, since acidic samples may prematurely shift the color of the reaction mix, even in non-infected saliva samples, thereby possibly rendering false positives. LAMP-based methods also use from four to six primer sets, and the design of LAMP primer sets is nontrivial as the primers may interact among themselves during amplification. This can promote non-specific amplifications that may also result in false positives[22,23].

In the present study, we demonstrate the straightforward use of microelectrodes to monitor, in real time, the evolution of the amplification of SARS-CoV-2 genetic material. This simple implementation renders a quantitative and label-free embodiment of the LAMP reaction (LF-LAMP) for amplification of any genetic sequence. Label free quantitative LAMP (LF-qLAMP) allows the continuous and online monitoring of the change in the electric potential of the reaction mix due to the release of hydrogen ions during amplification. We show that the progressions of the signal from truly positive samples (samples containing genetic material from SARS-CoV-2) and negative samples clearly differ in LF-qLAMP. Therefore, this method provides a straightforward way to distinguish pH shifts that are truly due to specific amplifications and are not false positives.

In this time of COVID-19, we also illustrate LF-qLAMP for the diagnostics of COVID-19 from saliva samples spiked with SARS-CoV-2 nucleic acids. Overall, we demonstrate the utility of extraction-free LF-LAMP for COVID-19 from samples containing artificial SARS-CoV-2 genetic material and from saliva samples containing artificial SARS-CoV-2 genetic material.

## Materials and methods

### Equipment specifications

We fabricated a simple and portable prototype for the isothermal incubation of samples and online monitoring of the LAMP amplification process using a commercial and low-cost pH sensor (Gaohau pH 0–14; available at Amazon.com; USA), a commercial pH electrode (inLab nano; Mettler Toledo, Switzerland), and an Arduino UNO® microprocessor (Arduino, Italy).

This *ad hoc* heating system was assembled with components acquired over the internet (amazon.com) and consisted of a 12V eliminator, a 10 mL vessel with a silicon lead, a heating mat (electric resistance), and an Arduino UNO–based proportional integral derivative (PID) temperature controller (fully described in the supplementary material). The Arduino-based PID controller is depicted in Figure 2A and Figure S1. The code used in our heating/amplification experiments has been made available as supplementary material (Supplementary file 1).

**Figure 1.**
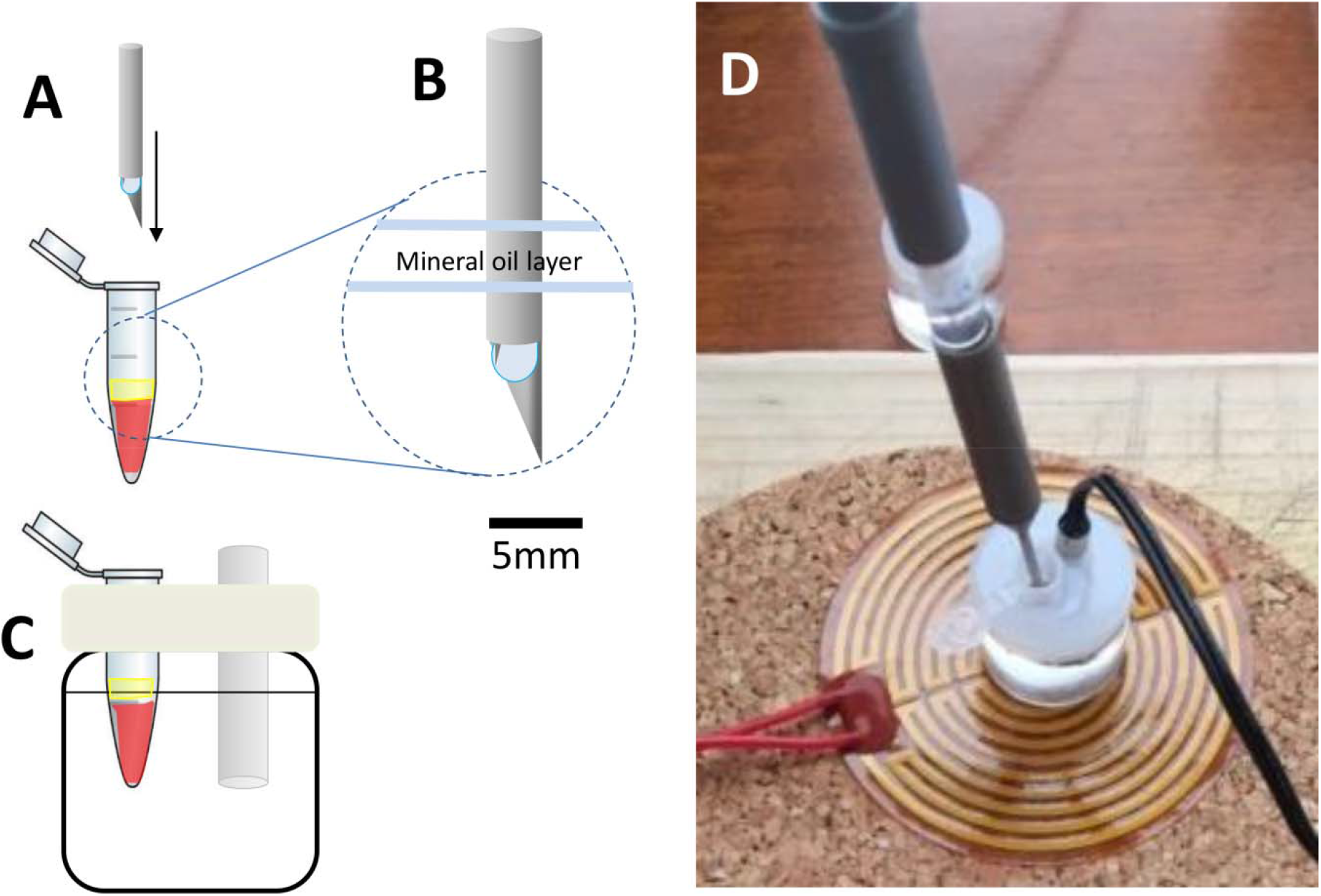
Experimental setup. (A) A micro pH electrode is inserted into the Loop-mediated Isothermal Amplification (LAMP) colorimetric reaction mix and (B) a thin layer of oil is added to avoid evaporation during incubation (C) in an oil bath. (D) Image of the actual system; a heating mat connected to an Arduino-based proportional integral derivative (PID) controller is used to control the temperature at 62.5 +/- 1.2 °C.

**Figure 2.**
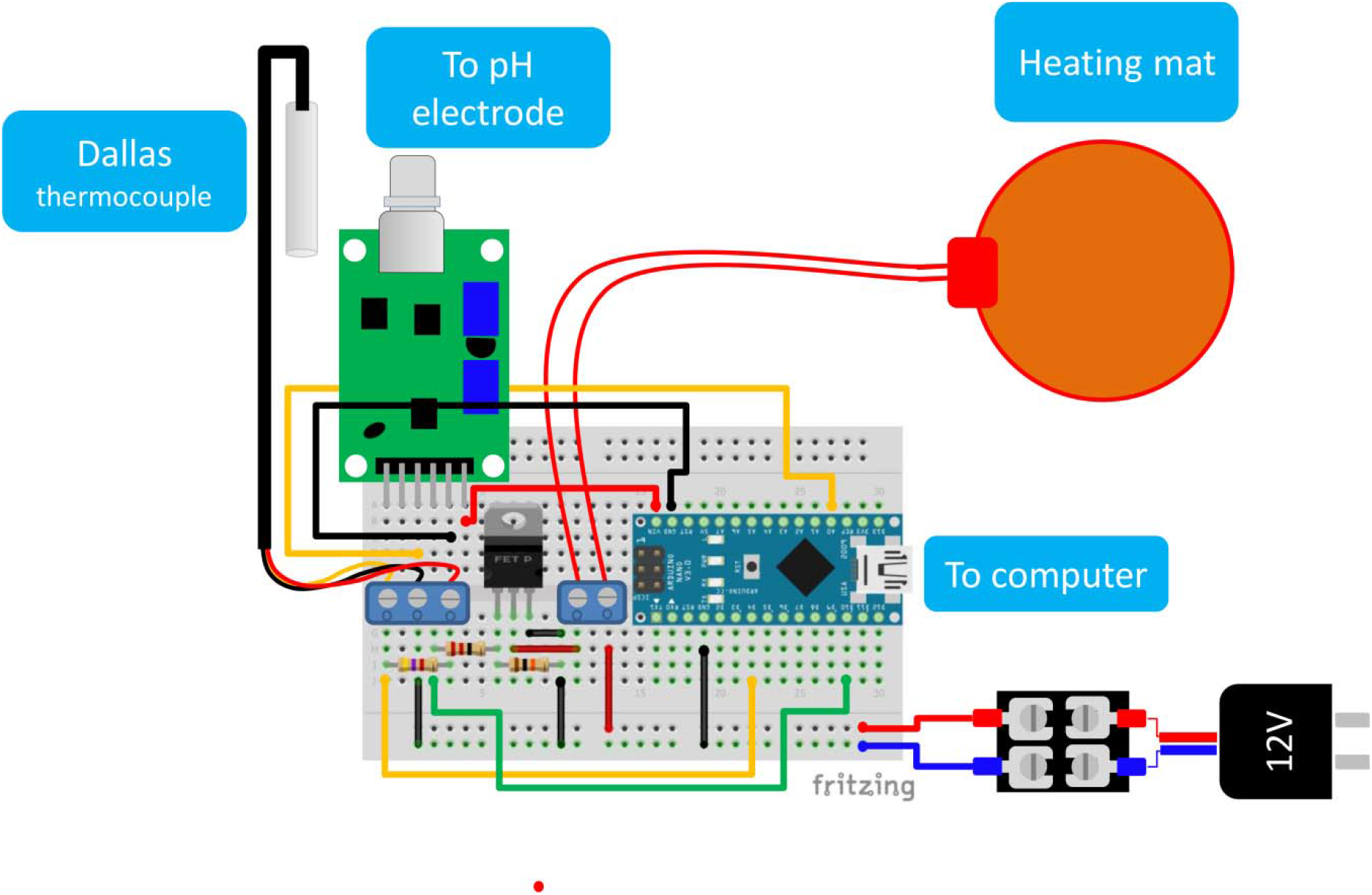
Schematic representation of the Arduino-based proportional integral derivative (PID) temperature controller and online electric potential monitoring system.

The evolution of the amplification process was followed using commercially available pH electrodes (inLab nano, from Mettler Toledo, Switzerland; and Lab Sen 241-3, from Aprea Instruments, available at Amazon.com). These electrodes were interfaced through an Arduino UNO microprocessor using a pH sensor module (Gaohou pH 0-14 detect sensor module, available at Amazon.com). The InLab microelectrode, with a tip diameter of 1.3 mm, can be easily placed inside a conventional 200 µL Eppendorf tube and enables the use of small reaction volumes (25–50 µL of reaction mix and sample). Alternatively, we have run online experiments using a lower cost electrode from Apera Instruments (∼150 USD) with a tip diameter of 3 mm, which can also be used with similar measuring performance (supplementary material; Figure S1) if a larger reaction volume is used (∼50 µL).

In our experiments, the insertion of the electrode within the reaction mix impedes closure of the Eppendorf tube during incubation. To avoid evaporation during heating, two drops of mineral oil were dispensed into each Eppendorf tube before electrode placement to allow the development of a thin lid layer of mineral oil at the liquid surface of the reaction mix. The Arduino code used in the experiments reported here are available as supplemental material (Supplemental file 1).

### Experiments with synthetic genetic material

We ran several series of experiments where samples containing different quantities of SARS-CoV-2 synthetic genetic material were analyzed using LF-qLAMP. These amplification experiments were conducted using the homemade system previously described for isothermal heating of the samples at 62.5 +/-1.5 °C and for online monitoring of the progression of the electric potential in the reaction mix during amplification.

Samples were prepared by adding 4 µL of a solution containing the DNA template to 250 µL Eppendorf tubes containing 25 µL WarmStart^®^ Colorimetric LAMP 2× Master Mix (DNA & RNA) from New England Biolabs (MA, USA), 18 µL nucleasease-free water from New England Biolabs (MA, USA), and 2 µL primer solution. Samples were prepared by adding different quantities of positive DNA templates (i.e., 0, 10, 100, 1,000, and 10,000 copies of the gene N from SARS-CoV-2; Integrated DNA Technologies, IA, USA). Negative amplification controls were prepared using the reaction mix, as previously described, but without primer addition.

### Preparation of saliva samples spiked with RNA extracts

A set of 5 saliva samples obtained from healthy volunteers was spiked with different quantities of SARS-CoV-2 synthetic genetic material. Saliva samples were also collected from a COVID-19(+) patient after obtaining written consent and in full compliance with the principles stated in the Helsinki Declaration. Every precaution has been taken to protect the privacy of sample donors and the confidentiality of their personal information. Our experimental protocols for saliva collection and saliva use in amplification was approved, as part of a wider study to collect and test saliva samples from human volunteers, by the Alfa Medical Research Committee in the resolution AMCCI-TECCOVID-001 on May 20th, 2020 (Alfa Medical Center, Research Committee; Monterrey, NL, México). Saliva samples were inactivated by heat treatment at 62.5 °C for 35 minutes prior to amplification.

### Amplification mix

We used WarmStart^®^ Colorimetric LAMP 2× Master Mix (DNA & RNA) from New England Biolabs (MA, USA), and followed the recommended protocol: 25 μL Readymix, 1.6 μM FIP primer, 1.6 μM BIP primer, 0.2 μM F3 primer, 0.2 μM B3 primer, 0.4 μM LF primer, 0.4 μM LB primer, 2 μL DNA template (∼ 625 to 2 × 10^5^ DNA copies), and nuclease-free water in a final reaction volume of 50 μL. This commercial mix contains phenol red as a pH indicator to reveal the pH shift occurring during LAMP amplification across the threshold of pH=6.8.

### Primers used

A sets of six LAMP primers, referred to here as the α-set, was designed in house using the LAMP primer design software Primer Explorer V5 (http://primerexplorer.jp/lampv5e/index.html). These LAMP primers were designed to target a region of the sequence of the SARS-CoV-2 N gene; specifically, they were based on the analysis of alignments of the SARS-CoV-2 N gene sequences using the Geneious software (Auckland, New Zealand).

RT-qPCR amplification experiments were conducted using one the primer sets recommended by the Centers for Disease Control (CDC) for the standard diagnostics of COVID-19 (i.e., the N1 assay). The sequences of our LAMP primers are presented in Table 1. The sequences of the PCR primers (N1) have been reported elsewhere [24,25].

**Table 1.**
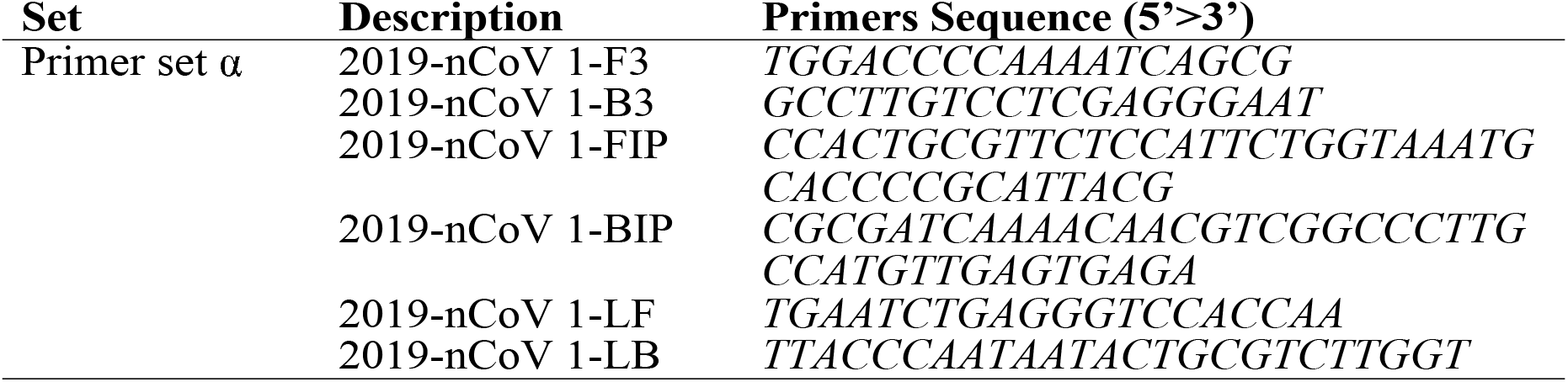
Sequences of the Loop-mediated Isothermal Amplification (LAMP) primers used for the detection of segments of genetic material encoding for the expression of the N protein of SARS-CoV-2.

### Amplification protocols

For all LAMP experiments, we performed isothermal heating at 62.5 +/-1.2 °C for 30 to 60 min.

## Results and discussion

### Rationale of the design and operation

Here, we have demonstrated the use a portable Arduino-based LAMP system as a label-free quantitative method (LF-qLAMP) for the reliable and fast identification of samples containing SARS-CoV-2 genetic material. We have introduced a simple and fully portable setup for the detection of viral genetic material using commercially available pH microelectrodes for real-time monitoring of the progression of LAMP reactions. Remarkably, this LAMP-based method is capable of discerning between positive and negative samples during the first three minutes of amplification.

The rationale behind the operation of this strategy is straightforward. In the weakly buffered reaction mix used for LAMP, the release of H_3_O^+^ ions inherent in the amplification process [26] can be measured in real time as a difference in the electric potential[27,28] or a pH change [29,30]. We fabricated a simple and portable prototype to accomplish two purposes: the isothermal incubation of samples and the online monitoring of the amplification process. To this aim, we used a commercial and low-cost pH sensor, a commercial pH electrode, and an Arduino UNO® microprocessor.

Figure 1 shows the different aspects of this prototype. The isothermal amplification reactions were conducted at 62.5 °C +/-1.5 °C in 200 µL Eppendorf tubes immersed in a mineral oil bath (Figure 1A) and containing a total reaction mix volume of 50 µL. The tip of a commercial pH microelectrode was immersed into the reaction mix for online monitoring of the progression of the electric potential (in mV) using an Arduino-based system. A simple Arduino code was used to enable the collection and transmission of data to the computer (Supplementary File S1); the difference in the electric potential (ΔP), as evaluated in the reaction solution, was continuously transmitted and recorded at a sampling rate of 1 Hz (i.e., 1 sampling point per second).

The stability of the electric potential readings is affected by temperature fluctuations around the set point. Therefore, adequate temperature control (62.5 +/-1.5 °C) is highly recommended for conclusive results. In our experiments, executed at home, the temperature within the water bath was controlled at 62.5 °C using the Arduino-based PID controller. For convenience, we have included all the information needed to build this portable and low-cost isothermal PID-incubator for Eppendorf tubes that enables full portability (Figure 1; Figure 2) of the system. Alternatively, the LAMP reaction can be incubated in any commercial thermo block or in a miniPCR® apparatus [31].

Figure 2 schematically shows the connections that should be made between the Arduino microcontroller, the low-cost pH sensor, and the rest of the components of the circuit that constitute the PID control system and the online electric potential monitor. Figure S1 (in supplementary material) shows an image of the actual experimental Arduino-based PID controller system.

### Characterization of the performance of the monitoring system

We evaluated the stability of the potential signal at the temperature set point and during temperature ramps. The electric potential is a strong function of the temperature (Figure 3A). For instance, the electric potential increased ∼90 mV per 10°C in our experiments in the range of 45 to 55 °C. By contrast, at isothermal conditions, the intrinsic variation of the electric potential was very stable; the variations were lower than 0.3% for the range of pH buffers that we assayed.

**Figure 3.**
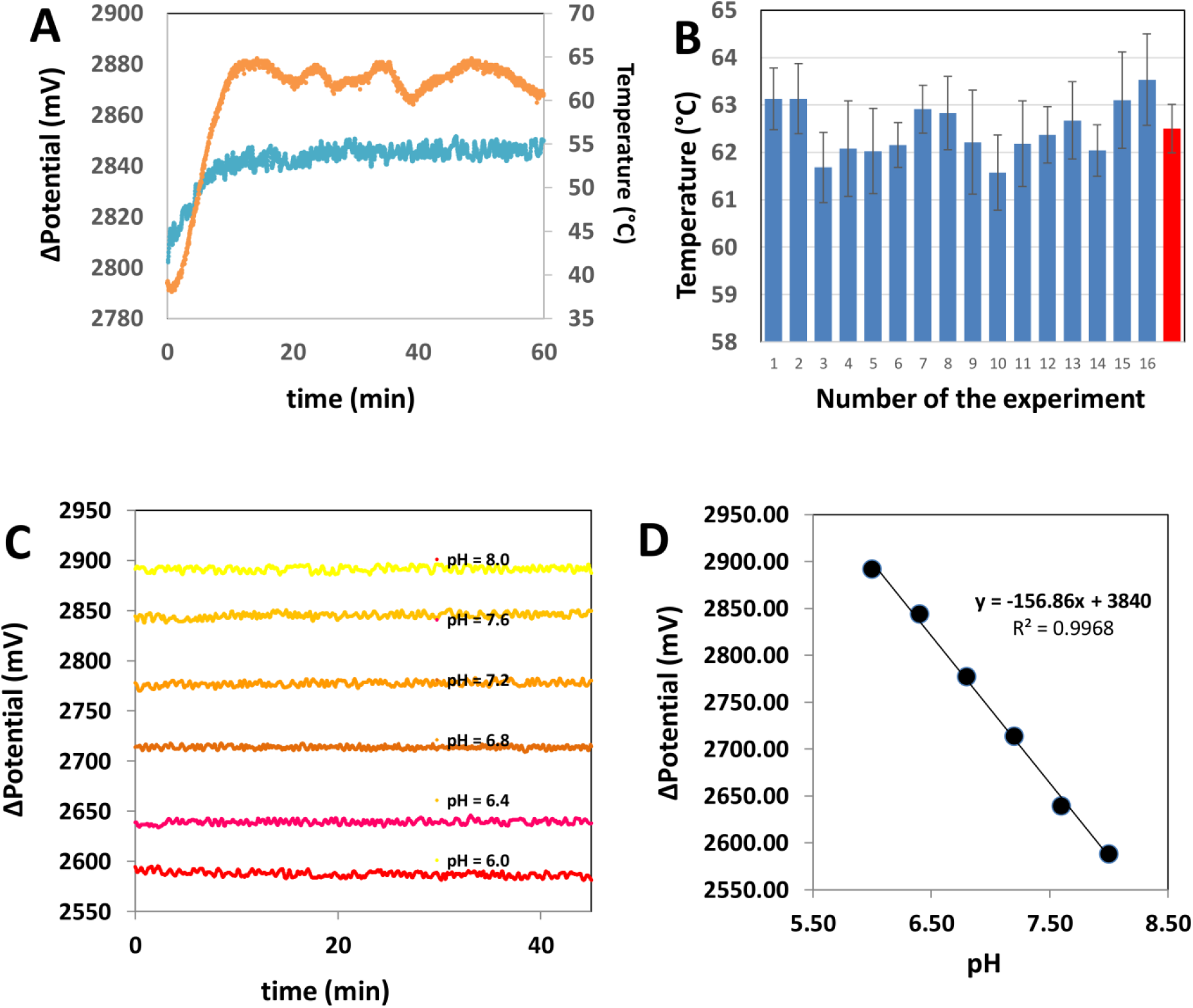
Basic characterization of the electric potential monitoring system and the Arduino-based proportional integral derivative (PID) controller. (A) The potential signal is stable at different pH values. (B) A linear dependence between pH and potential differential was observed in the entire range of pH values relevant to the Loop-mediated Isothermal Amplification (LAMP) reaction. (C) The potential differential is stable at isothermal conditions. (D) The Arduino-based PID controller is able to maintain the temperature value around the set point value (62.5 +/- 0.5° C) in a set of 16 independent experiments. The red bar indicates the average temperature and standard deviations of this set of 16 independent experiments.

We also analyzed the ability of this simple PID controller to function for extended incubation periods (i.e., 60 minutes) and under different room temperatures (Figure 3B). Overall, we found that this simple incubator system is sufficiently robust to maintain the temperature at the set point (average temperature of 62.5 °C; average standard deviation of 0.5 °C) regardless of the room temperature (i.e., in a range between 15 and 25 °C). As we will show, this level of control is sufficient to obtain reliable and reproducible results of determination of the electric potential in solutions with different pH values and during LAMP reactions.

In a first set of experiments, we evaluated the robustness and consistency of the characteristics of the electrode readings at different pH values. Figure 3A shows representative readings associated with the immersion of the microelectrode in buffer solution of different pH values (i.e., pH 6.0, 6.4, 6.8, 7.2, 7.6, and 8.0).

The standard deviation associated with the readings within this window of pH values ranges from 1.38 and 6.91 mV (Table S1). For example, the average electric potential measured for a buffer solution at pH 6.8 (which is approximately the threshold value of phenol red) was 2777 mV and exhibited a standard deviation of +/-1.38 mV.

A buffer solution at pH 8.0 (which is similar to the pH value of the LAMP buffer used) had an electric potential of 2587.58 mV with a standard deviation of +/-6.21 mV. Overall, we observed coefficients of variance lower than 0.3% for all the experiments conducted in the absence of amplification reactions. In all these cases, the electric potential remains essentially stable during incubation periods of 60 minutes. This indicates that the inherent error (i.e., the intrinsic fluctuation associated with the potential signal at the experimental conditions) is in the range of +/-12.3 mV (i.e., the maximum standard deviations are lower than 7 mV). We also evaluated the stability of the signal reported by the microelectrode when immersed in the buffer of the reaction mix. Our results also suggest that the potential readings obtained using this simple sensing system are robust and steady, with intrinsic coefficients of variance lower than 1%.

### Discrimination between positive and negative samples

Using the simple Arduino-based system that we have described, we incubated a series of LAMP reactions and monitored the progress of the amplification reaction by measuring the electric potential in the reaction mix in real time. We successfully discriminated between COVID-19(+) and COVID-19(-) samples in a set of synthetic samples and in a set of saliva samples spiked with synthetic SARS-CoV-2 genetic material.

We first characterized the baseline corresponding to negative samples, where no primers were added (supplementary material; Figure S2), As expected, the signal associated with these samples was relatively steady (average standard deviations ∼10.7; n=3). We then monitored the evolution of the LAMP reactions in the negative samples containing LAMP reaction mix and primers, but no synthetic SARS-CoV-2 genetic material.

One of the limitations of colorimetric LAMP-based methods based on the phenol red color shift is that some degree of color change exists in samples containing no template (see also ref [27]). This has been associated to the occurrence of non-specific amplification induced by the interaction among the LAMP primers [23,32], which often serve as templates for off-target polymerizations. In general, non-specific amplification is believed to be the main reason for false positives in LAMP colorimetric reactions.

Figure 4 A shows the typical profile of the progression of the electric potential in negative samples (with primers, reaction mix, but no SARS-CoV-2 template). Four representative replicates are shown. The shape of the progression of the electric potential in the negative samples is highly consistent. Figure 4B presents a normalization of the four representative curves shown in Figure 4A. The curves were normalized by subtracting the initial potential value for each curve. In doing so, all curves practically collapse into an invariant shape.

**Figure 4.**
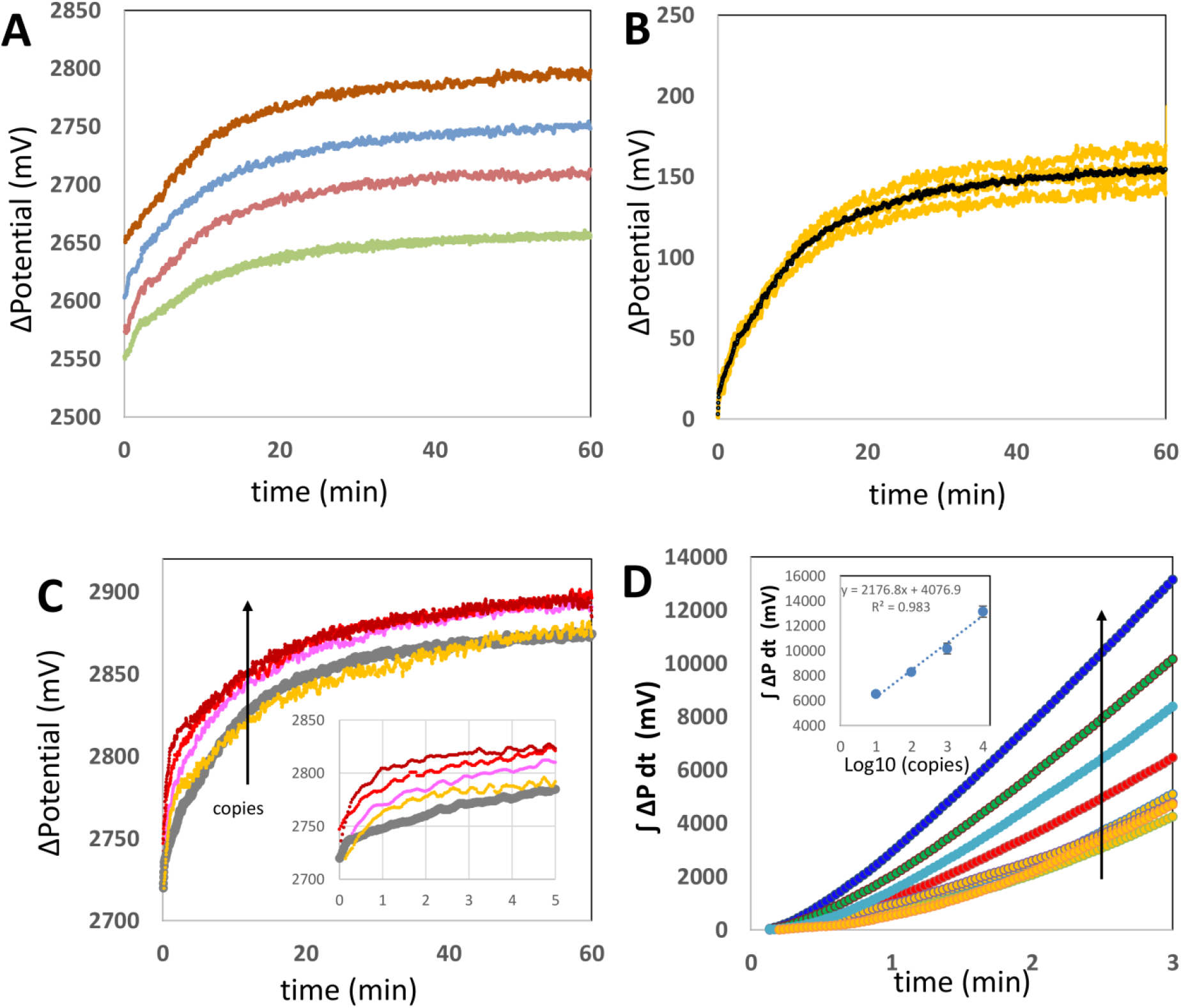
Discrimination between SARS-CoV-2 positive and negative samples using online electric potential measurements. (A) Measurement of four different negative samples (i.e., samples without SARS-CoV-2 genetic material, but containing reaction mix and primers). (B) Normalized signal of a set of four different negative samples indicated with yellow lines. Values were normalized by subtracting the initial potential value of each curve. The black line depicts the average of the four yellow trajectories and is characteristic of the progression of the Loop-mediated Isothermal Amplification (LAMP) reaction observed in negative samples. (C) Comparison of the online potential signal in samples containing 10,000 (magenta), 1000 (red), 100 (pink), and 10 (yellow) copies of synthetic SARS-CoV-2 genetic material. The average of four negative samples is depicted in gray. The inset shows a close-up of the first five minutes of the amplification reaction. (D) Plot of the integral of the differential of potential with respect to time for the first 3 minutes of amplification in samples containing different copy numbers of synthetic SARS-CoV-2 genetic material: 10,000 (dark blue), 1000 (brown), 100 (light blue), and 10 (red) copies of synthetic SARS-CoV-2 genetic material. Trends associated with negative samples are presented in blue, orange, green, and yellow.

The black line in Figure 4B depicts the average of the four curves corresponding to independently run negative samples. This profile, characteristic of negative samples, is distinguishable from that inherent to the specific amplification observed in samples that do contain SARS-CoV-2 genetic material. Figure 4C shows the trajectories of the electric potential associated with positive samples containing different quantities of SARS-CoV-2 synthetic genetic material (color curves).

The average progression of the electric potential in negative samples (from Figure 4B) is indicated by the gray line. The potential exhibits a distinctive, practically immediate, and sharp increase at the incubation conditions that are associated with a high initial rate of amplification. The initial slope of the potential curve is steeper in positive samples than in negative samples. However, a high sampling rate is required in the monitoring device for a clear identification of this steep initial slope. In our experiments, we achieved the resolution needed to identify the sharp slope associated with samples that contained SARS-CoV-2 genetic material by sampling at least once per second (the Arduino code is available in supplementary material; File S1). Lower sampling rates (e.g., 0.2 Hz) do not serve the purpose of clearly resolving the initial slope of the reaction in positive samples (supplementary material; Figure S3). We also observed that the evolution of each curve is consistent with the copy load in the positive samples, so that higher copy numbers originate curves with a higher initial slope than is observed for curves associated with lower loads. Since the initial rate of amplification is remarkably high in positive samples (> 25 to 75 mV s^-1^), the differences between slopes can be clearly established only if high sampling rates (∼ 1 sample s^-1^: 1 Hz) are imposed.

In general, discrimination in positive samples between negative samples and positive samples with medium to high viral loads (i.e., at least 100 copies) by comparison of the potential trajectories. However, samples containing a low copy number (i.e., less than 10 gene copies) cannot be distinguished from negative samples by only comparing trajectories (Figure 4C). The inset in Figure 4C zooms in on the first five minutes of the amplification reaction and shows that the behavior of the trajectory of the amplification reaction in positive and negative samples can be better discriminated at this first stage of the amplification. For instance, the value of the area under the curve or of ΔP versus t is distinguishably higher in positive samples than in negative samples during the first stage of the amplification. Here, we define IP3, the integral of the potential with respect to time for the first 3 minutes of reaction, as an indicator of specific amplification of genetic material in the LAMP reactions (equation 1).

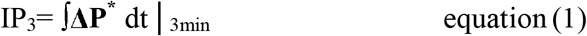

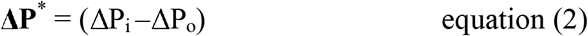

Here, ΔP_i_ is the value of electric potential measured at every sampling point and ΔP _o_ is the value of electric potential at the beginning of the experiment. Therefore **ΔP** ^**\***^is the increment in potential measured at each sampling event (i.e., in our case, every second) with respect to the initial value of electric potential in the reaction mix at the initial point of the amplification. Equation 3 provides a step by step approximation of IP_3_.

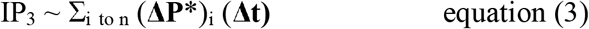

Here, n is the number of sampling points within the time frame from 0 to 3 minutes. Indeed, we found that IP_3_ is a reliable and robust predictor that enables the consistent and rapid identification of samples containing (or not containing) SARS-CoV-2 genetic material. Figure 4C shows the evolution of the integral of IP_3_ for the first 3 minutes of a set of LAMP reactions. Progressions corresponding to samples added with different quantities of synthetic genetic material (i.e., 10, 100, 1000, and 10000 copies of the N gene of SARS-CoV-2) were compared versus three independent repeats of negative samples (i.e., samples without SARS-CoV-2 genetic material). Negative samples (plotted in yellow) exhibit integrals significantly lower than those calculated from the analysis of positive samples. In addition, a linear correlation can be established between the logarithm of the copy number and the value of the integral (Figure 4D; inset). These results suggest that IP_3_ is a reliable and quantitative indicator of the specific amplification of genetic material in simple samples.

### qLAMP in saliva samples

We then extended the use of this embodiment of qLAMP to the identification of SARS-CoV-2 genetic material in saliva samples. As already mentioned, colorimetric LAMP using phenol red has recently received attention as a cost-effective diagnostic method that enables discrimination between COVID(+) and COVID(-) samples by naked eye inspection and is therefore independent of the use of costly PCR instruments[11,33].

However, colorimetric LAMP exhibits its own important limitations, and its direct implementation in saliva samples is technically challenging. Human saliva exhibits a wide range of pH values[34,35]; therefore, even small volumes of saliva may significantly affect the initial pH of the reaction mix in LAMP-based methods. Acidic saliva samples may induce a premature shift in the pH (and the color) that is not associated with amplification and will eventually lead to a false positive diagnosis. The direct and real-time reading of electric potential throughout the amplification offers a solution to some of the limitations of final-point colorimetric LAMp methods in the context of the analysis of saliva samples.

Figure 5A shows the results of the progression of LAMP amplification in saliva samples with or without added synthetic genetic material in different quantities (i.e., in the range of 10 to 10,000 copies of the N gene of SARS-CoV-2). In this experimental set, the reaction mix contained 3–4 µL of real undiluted saliva (volume defined based in ref [15]) from SARS-CoV-2 negative volunteers (as determined by end-point colorimetric LAMP). As before, the amplification curves that corresponded to samples spiked with SARS-CoV-2 genetic material exhibited a steep slope that was immediately observable upon initiation of the incubation. The typical smoother progression associated with negative samples (Figure 4B) was also observed in negative saliva samples. In Figure 5B, we have plotted the results from a set of experiments in which the IP_3_ values for saliva samples with or without SARS-CoV-2 genetic material were calculated. The IP_3_ values were clearly higher in positive samples than in negative samples. The positive samples with low to medium SARS-CoV-2 gene copy loads were also clearly distinguishable from negative samples.

**Figure 5.**
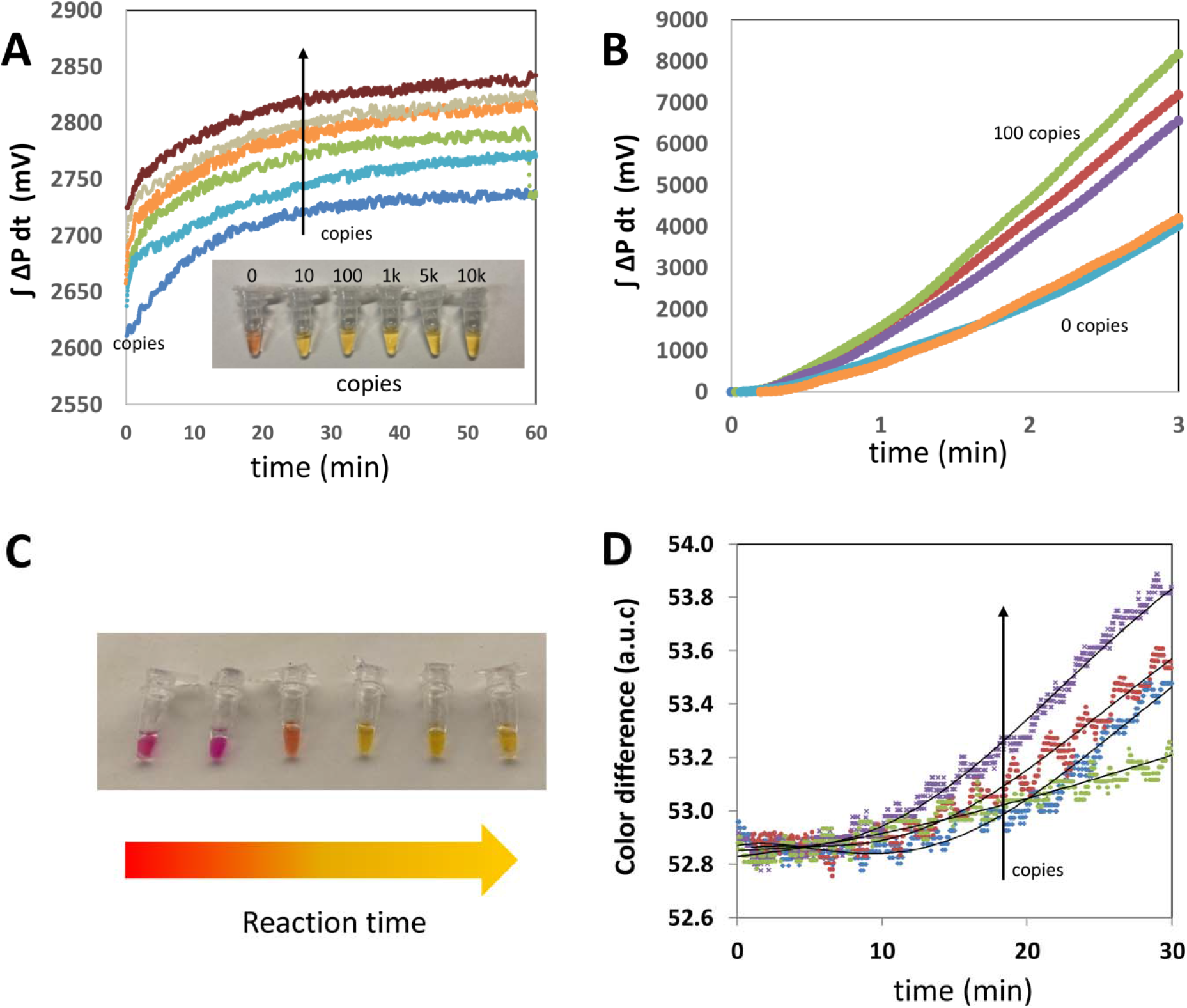
Discrimination between SARS-CoV-2 positive and negative saliva samples using online electric potential measurements. (A) Results from the online evaluation of electric potential in a saliva sample from a volunteer diagnosed as SARS-CoV-2 negative (dark blue curve) and a set of five saliva samples added with 10 (blue line), 100 (green line), 1000 (orange curve), 5000 (beige curve), and 10000 copies (brown curve) of SARS-CoV-2 genetic material. (B) Plot of the integral of the differential of potential with respect to time for the first 3 minutes of amplification in saliva samples containing different copy numbers of synthetic SARS-CoV-2 genetic material in the range of 10–100 copies (purple, green and red curves) or not containing SARS-CoV-2 genetic material (orange and blue curves). The blue curve corresponds to the analysis of an actual sample from a symptomatic patient later confirmed as SARS-CoV-2 negative by RT-qPCR. (C) Typical color progression (from red-magenta to crisp yellow in a positive sample) through colorimetric Loop-mediated Isothermal Amplification (LAMP). (D) Evolution of the difference in color (with respect to the initial red-magenta color) in positive samples containing 0 (green), 10 (blue), 100 (red), and 1000 (purple) added synthetic copies of SARS-CoV-2 genetic material.

The IP_3_ value is an indicator of the extent of specific amplification that is somehow analogous to the progression of the change of color (from red-magenta to crisp yellow; Figure 5C) during the amplification in colorimetric LAMP. Indeed, the use of a commercial color sensor allows the quantitative analysis of the process of the color change during colorimetric LAMP[36] mediated by phenol red, as we and others have shown previously [11,37] (supplementary material; Figure S4). For comparison, we monitored online the change in color associated with the process of amplification in samples that contained different quantities of SARS-CoV-2 genetic material (Figure 5D).

Note that the shape of the curves that describes the change in color and the change in potential are similar. This is expected. Both processes (i.e., the change in electric potential in the reaction mix and the development of color) are cumulative processes directly related to the release of protons during amplification. However, the time scale at which color and electric potential evolve differs quite drastically. Changes in color are observable at a much slower rate—positive and negative samples can be discriminated due to their color evolution within the first 30 minutes of reaction (supplementary material; Figure S4 A and C). Remarkably, the evolution of the IP_3_ indicator introduced here enables a reliable discrimination between positive and negative samples within the initial 3 minutes of the reaction.

## Potential relevance and limitations

The result presented here may be highly relevant in practical terms. The typical concentration of gene copies of SARS-CoV-2 in the saliva of positive subjects (even asymptomatic ones) is generally higher than 10^5^ mL^-1^ (10^3^ µL^-1^). In the analysis presented here, 3–4 µL^-1^ of undiluted saliva can be added without observing inhibitory effects in terms of the extent of the amplification. Therefore, copy number values greater than 10^3^ per assay can be expected. Based on our experiments, the limit of detection/discrimination of this method, assisted by online monitoring of the electric potential of the solution during amplification, is two orders of magnitude below this (i.e., 10–100 SARS-CoV-2 copies per assay). This suggests that this method could render reliable diagnostic results for diagnosis of COVID-19 in unextracted (undiluted or diluted) saliva samples that have even moderate or low viral loads. This hypothesis remains to be tested by the diagnostic evaluation of saliva samples from actual patients and at this point should be taken as an inference.

Several limitations of the methodology presented here should also be noted. The successful performance of this diagnostic strategy is centered on the ability to sample the electric potential (or pH) of the reaction mix frequently (the authors recommend at least at 1.0 Hz) during the initial stage of the amplification reaction (i.e., at least during the initial three minutes). Therefore, this requires an electrode with a sufficiently fast response and a sensor capable of operating at a sufficiently high sampling rate (at least one pH measurement per second). In addition, equilibration of the electrode at the typical initial pH (or potential) of the LAMP reaction mix is desirable. Failure to do this may hinder the resolution of the readings during the first seconds of the amplification due to a lag in the response or the adjustment of the sensor to the pH conditions (supplementary material; Figure S3).

In the current developmental state of our diagnostic device, we depend on the use of commercial pH microelectrodes that are relatively expensive. For instance, we estimate that the cost of development a diagnostic unit capable of running 8 reactions simultaneously in our lab would be on the order of 10,000 USD. In addition, the use of glass or membrane-based pH microelectrodes may be a source of contamination of samples with residual DNA (from a previous positive amplification). Protocols should be established that can effectively remove residual nucleic acids after the completion of an amplification experiment from the electrode. The development of disposable and low-cost pH microelectrodes should be a priority for enabling the cost-effective use of this strategy to intensify the ability to perform label-free and portable/mobile molecular diagnostics of SARS-CoV-2 and other pathogens.

## Conclusions and outlook

Preparedness to face infectious diseases is the key to buffering the spread of epidemics and minimizing the number of human deaths. Extraction-free and portable diagnostics from saliva will greatly enable the intensification of testing efforts in the context of pandemic COVID-19 and other infectious diseases.

Here, we have introduced a strategy for reliable and consistent identification of samples containing SARS-CoV-2 genetic material. This strategy relies on the real-time monitoring of the LAMP reaction in the weakly buffered reaction mix commonly used for colorimetric LAMP. The continuous release of protons associated with the incorporation of bases into the nascent chains of DNA during amplification is measured online with a pH microelectrode that is immersed in the reaction mix incubated at 62.5 °C. We also showed that proper isothermal control and online determination of changes in potential (or pH) in the LAMP reaction mix can be appropriately accomplished using a simple Arduino-based device.

In addition, we demonstrate that the area under the curve that describes the time evolution of the values of the electric potential of the reaction mix during the first three minutes of the amplification (i.e., defined here as IP_3_) is a reliable predictor of the extent of specific amplification occurring during the LAMP reactions. The evaluation of IP_3_ enables the consistent discrimination between samples containing or not containing SARS-CoV-2 genetic material. We also demonstrated the feasibility of using this portable, fast, and reliable diagnostic strategy with saliva samples containing different loads of SARS-CoV-2 genetic material.

Recent reports have shown that the performance of LAMP is greatly influenced by the reaction conditions and the reagents used (i.e., primers sets, retro-transcriptases or proteases, additives, and fluorescent dyes, among others) [17,38]. The label-free qLAMP platform presented here may be highly useful in the selection of reaction reagents (i.e., alternative polymerases, additives to enhance the amplification, or primer sets) or optimization of LAMP conditions (i.e., temperatures and concentrations). The mere comparison of the curves of evolution of protons with time under different reaction conditions provides a direct way to evaluate the effect of any modifications in the protocol on the overall performance of the amplification.

The release of protons during amplification, which is the underlying process that we are monitoring, is inherent in all nucleic acid amplification methods. Therefore, the methodology of real-time monitoring of the change in electric potential during amplification is fully translatable to other amplification schemes (i.e., qPCR and RPA, among others).

## Data Availability

All data relevant to the writing of this article is available upon request.

## Acknowledgments

EGG and SB acknowledge funding from doctoral scholarship provided by CONACyT (Consejo Nacional de Ciencia y Tecnología, México). GTdS and MMA acknowledge the institutional funding received from Tecnológico de Monterrey (Grant 002EICIS01 and Novus 2019) and CONACyT (Sistema Nacional de Investigadores; 66839 and 26048). The authors acknowledge the funding provided by the Federico Baur Endowed Chair in Nanotechnology (0020240I03). The authors gratefully acknowledge funding from Fundación-FEMSA and from an alumnus of Tecnológico de Monterrey who prefers to remain anonymous.

## CRediT author statement

**Mario Moisés Alvarez:** Conceptualization, Methodology, Investigation, Formal Analysis, Resources, Software, Data curation, Writing - original draft, Writing-Reviewing & Editing, Supervision, Project Administration, Funding acquisition; **Sergio Bravo-González**: Methodology, Investigation, Data curation, Software; **Everardo González-González:** Methodology, Investigation, Resources; **Grissel Trujillo-de Santiago:** Conceptualization, Methodology, Resources, Writing-Reviewing & Editing, Supervision, Project Administration, Funding acquisition.

## Competing interest

The authors declare no competing interests.

## Notes

### Competing Interest Statement

We are currently in the process of signing a collaboration agreement with Aim Brands, Llc.

### Funding Statement

We acknowledge funding from Tecnologico de Monterrey (Novus; Office of Technology Transfer - COVID-19 Emergency Funding). We are grateful and acknowledge support from Fundacion-FEMSA and form an altruist Tecnologico de Monterrey alumni who prefers to remain anonymous.

### Author Declarations

The experimental protocol was approved on May 20th, 2020 by a named institutional committee (Alfa Medical Center, Research Committee; resolution AMCCI-TECCOVID-001).

## References

[1] Home - Johns Hopkins Coronavirus Resource Center, (n.d.). https://coronavirus.jhu.edu/ (accessed September 10, 2020).

[2] M.N. Esbin, O.N. Whitney, S. Chong, A. Maurer, X. Darzacq, R. Tjian, Overcoming the bottleneck to widespread testing: A rapid review of nucleic acid testing approaches for COVID-19 detection, RNA. 26 (2020) 771–783. doi:10.1261/rna.076232.120.

[3] N. Younes, D.W. Al-Sadeq, H. Al-Jighefee, S. Younes, O. Al-Jamal, H.I. Daas, H.M. Yassine, G.K. Nasrallah, Challenges in Laboratory Diagnosis of the Novel Coronavirus SARS-CoV-2, Viruses. 12 (2020) 582. doi:10.3390/v12060582.

[4] M.P. Cheng, J. Papenburg, M. Desjardins, S. Kanjilal, C. Quach, M. Libman, S. Dittrich, C.P. Yansouni, Diagnostic Testing for Severe Acute Respiratory Syndrome-Related Coronavirus 2: A Narrative Review, Ann. Intern. Med. 172 (2020) 726–734. doi:10.7326/M20-1301.

[5] C.B.F. Vogels, A.F. Brito, A.L. Wyllie, J.R. Fauver, I.M. Ott, C.C. Kalinich, M.E. Petrone, A. Casanovas-Massana, M. Catherine Muenker, A.J. Moore, J. Klein, P. Lu, A. Lu-Culligan, X. Jiang, D.J. Kim, E. Kudo, T. Mao, M. Moriyama, J.E. Oh, A. Park, J. Silva, E. Song, T. Takahashi, M. Taura, M. Tokuyama, A. Venkataraman, O. El Weizman, P. Wong, Y. Yang, N.R. Cheemarla, E.B. White, S. Lapidus, R. Earnest, B. Geng, P. Vijayakumar, C. Odio, J. Fournier, S. Bermejo, S. Farhadian, C.S. Dela Cruz, A. Iwasaki, A.I. Ko, M.L. Landry, E.F. Foxman, N.D. Grubaugh, Analytical sensitivity and efficiency comparisons of SARS-CoV-2 RT–qPCR primer–probe sets, Nat. Microbiol. 5 (2020) 1299–1305. doi:10.1038/s41564-020-0761-6.

[6] J.M. Sharfstein, S.J. Becker, M.M. Mello, Diagnostic Testing for the Novel Coronavirus, JAMA - J. Am. Med. Assoc. 323 (2020) 1437–1438. doi:10.1001/jama.2020.3864.

[7] A.K. Giri, D.R. Rana, Charting the challenges behind the testing of COVID-19 in developing countries: Nepal as a case study, Biosaf. Heal. 2 (2020) 53–56. doi:10.1016/j.bsheal.2020.05.002.

[8] L. Yu, S. Wu, X. Hao, X. Dong, L. Mao, V. Pelechano, W.-H. Chen, X. Yin, Rapid Detection of COVID-19 Coronavirus Using a Reverse Transcriptional Loop-Mediated Isothermal Amplification (RT-LAMP) Diagnostic Platform, Clin. Chem. 66 (2020) 975–977. doi:10.1093/clinchem/hvaa102.

[9] L.E. Lamb, S.N. Bartolone, E. Ward, M.B. Chancellor, Rapid Detection of Novel Coronavirus (COVID19) by Reverse Transcription-Loop-Mediated Isothermal Amplification, SSRN Electron. J. (2020). doi:10.2139/ssrn.3539654.

[10] Y. Zhang, N. Odiwuor, J. Xiong, L. Sun, R.O. Nyaruaba, H. Wei, N.A. Tanner, Rapid Molecular Detection of SARS-CoV-2 (COVID-19) Virus RNA Using Colorimetric LAMP, MedRxiv. 2 (2020) 2020.02.26.20028373. doi:10.1101/2020.02.26.20028373.

[11] E. González-González, I.M. Lara-Mayorga, I.P. Rodríguez-Sánchez, Y.S. Zhang, S.O. Martínez-Chapa, G.T. Santiago, M.M. Alvarez, Colorimetric loop-mediated isothermal amplification (LAMP) for cost-effective and quantitative detection of SARS-CoV-2: the change in color in LAMP-based assays quantitatively correlates with viral copy number, Anal. Methods. 13 (2021) 169–178. doi:10.1039/d0ay01658f.

[12] J. Rodriguez-Manzano, K. Malpartida-Cardenas, N. Moser, I. Pennisi, M. Cavuto, L. Miglietta, A. Moniri, R. Penn, G. Satta, P. Randell, F. Davies, F. Bolt, W. Barclay, A. Holmes, P. Georgiou, Handheld Point-of-Care System for Rapid Detection of SARS-CoV-2 Extracted RNA in under 20 min, ACS Cent. Sci. (2021). doi:10.1021/acscentsci.0c01288.

[13] X. Zhu, X. Wang, L. Han, T. Chen, L. Wang, H. Li, S. Li, L. He, X. Fu, S. Chen, M. Xing, H. Chen, Y. Wang, Multiplex reverse transcription loop-mediated isothermal amplification combined with nanoparticle-based lateral flow biosensor for the diagnosis of COVID-19, Biosens. Bioelectron. 166 (2020) 112437. doi:10.1016/j.bios.2020.112437.

[14] N. Toppings, A. Mohon, Y. Lee, H. Kumar, D. Lee, R. Kapoor, G. Singh, L. Oberding, O. Abdullah, K. Kim, B. Berenger, D. Pillai, Saliva-Dry LAMP: A Rapid Near-Patient Detection System for SARS-CoV-2, (n.d.). doi:10.21203/rs.3.rs-141322/v1.

[15] M.A. Lalli, J.S. Langmade, X. Chen, C.C. Fronick, C.S. Sawyer, L.C. Burcea, M.N. Wilkinson, R.S. Fulton, M. Heinz, W.J. Buchser, R.D. Head, R.D. Mitra, J. Milbrandt, Rapid and Extraction-Free Detection of SARS-CoV-2 from Saliva by Colorimetric Reverse-Transcription Loop-Mediated Isothermal Amplification, Clin. Chem. 67 (2021) 415–424. doi:10.1093/clinchem/hvaa267.

[16] B.A. Rabe, C. Cepko, SARS-CoV-2 detection using isothermal amplification and a rapid, inexpensive protocol for sample inactivation and purification, Proc. Natl. Acad. Sci. U. S. A. 117 (2020) 24450–24458. doi:10.1073/pnas.2011221117.

[17] A. Alekseenko, D. Barrett, Y. Pareja-Sanchez, R.J. Howard, E. Strandback, H. Ampah-Korsah, U. Rovšnik, S. Zuniga-Veliz, A. Klenov, J. Malloo, S. Ye, X. Liu, B. Reinius, S.J. Elsässer, T. Nyman, G. Sandh, X. Yin, V. Pelechano, Direct detection of SARS-CoV-2 using non-commercial RT-LAMP reagents on heat-inactivated samples, Sci. Rep. 11 (2021) 1820. doi:10.1038/s41598-020-80352-8.

[18] N. L’Helgouach, P. Champigneux, F.S. Schneider, L. Molina, J. Espeut, M. Alali, J. Baptiste, L. Cardeur, B. Dubuc, V. Foulongne, F. Galtier, A. Makinson, G. Marin, M.C. Picot, A. Prieux-Lejeune, M. Quenot, F.C. Robles, N. Salvetat, D. Vetter, J. Reynes, F. Molina, EasyCOV: LAMP based rapid detection of SARS□CoV□2 in saliva, MedRxiv. (2020) 2020.05.30.20117291. doi:10.1101/2020.05.30.20117291.

[19] E. González-González, I.M. Lara-Mayorga, I.P. Rodríguez-Sánchez, Y.S. Zhang, S.O. Martinez-Chapa, G. Trujillo de Santiago, M.M. Alvarez, Colorimetric Loop-mediated Isothermal Amplification (LAMP) for cost-effective and quantitative detection of SARS-CoV-2: The change in color in LAMP-based assays quantitatively correlates with viral copy number., Anal. Methods. (2020). doi:10.1039/D0AY01658F.

[20] N.A. Tanner, Y. Zhang, T.C. Evans, Visual detection of isothermal nucleic acid amplification using pH-sensitive dyes, Biotechniques. 58 (2015) 59–68. doi:10.2144/000114253.

[21] M.J. Kellner, J.J. Ross, J. Schnabl, M.P.S. Dekens, R. Heinen, I. Grishkovskaya, B. Bauer, J. Stadlmann, L. Menéndez-Arias, R. Fritsche-Polanz, M. Traugott, T. Seitz, A. Zoufaly, M. Födinger, C. Wenisch, J. Zuber, A. Pauli, J. Brennecke, S. Ameres, B. Bauer, N. Beer, K. Bergauer, W. Binder, C. Blaukopf, B. Bochev, J. Brennecke, S. Brinnich, A. Bundalo, M. Busslinger, A. Bykov, T. Clausen, L. Cochella, G. de Vries, M. Dekens, D. Drechsel, Z. Dzupinkova, M. Eckmann-Mader, U. Elling, M. Fellner, T. Fellner, L. Fin, B.V. Gapp, G. Grabmann, I. Grishkovskaya, A. Hagelkruys, B. Hajdusits, D. Haselbach, R. Heinen, L. Hill, D. Hoffmann, S. Horer, H. Isemann, R. Kalis, M. Kellner, J. Kley, T. Köcher, A. Köhler, D. Kordic, C. Krauditsch, S. Kula, R. Latham, M.C. Leitner, T. Leonard, D. Lindenhofer, R.A. Manzenreither, K. Mechtler, A. Meinhart, S. Mereiter, T. Micheler, P. Moeseneder, S. Nimpf, M. Nordborg, E. Ogris, M. Pagani, A. Pauli, J.M. Peters, P. Pjevac, C. Plaschka, M. Rath, D. Reumann, S. Rieser, M. Rocha-Hasler, A. Rodriguez, J.J. Ross, H. Scheuch, K. Schindler, C. Schmidt, H. Schmidt, J. Schnabl, S. Schüchner, T. Schwickert, A. Sommer, J. Stadlmann, A. Stark, P. Steinlein, S. Strobl, Q. Sun, W. Tang, L. Trübestein, C. Umkehrer, S. Urmosi-Incze, K. Uzunova, G. Versteeg, A. Vogt, V. Vogt, M. Wagner, M. Weissenboeck, B. Werner, R. Yelagandula, J. Zuber, A rapid, highly sensitive and open-access SARS-CoV-2 detection assay for laboratory and home testing, BioRxiv. (2020). doi:10.1101/2020.06.23.166397.

[22] J.C. Rolando, E. Jue, J.T. Barlow, R.F. Ismagilov, Real-time kinetics and high-resolution melt curves in single-molecule digital LAMP to differentiate and study specific and non-specific amplification, Nucleic Acids Res. 48 (2021) 42. doi:10.1093/NAR/GKAA099.

[23] X. Gao, B. Sun, Y. Guan, Pullulan reduces the non-specific amplification of loop-mediated isothermal amplification (LAMP), Anal. Bioanal. Chem. 411 (2019) 1211–1218. doi:10.1007/s00216-018-1552-2.

[24] E. Gonzalez-Gonzalez, G.T. Santiago, I.M. Lara-Mayorga, S.O. Martinez-Chapa, M.M. Alvarez, Portable and accurate diagnostics for COVID-19: Combined use of the miniPCR thermocycler and a well-plate reader for SARS-Co2 virus detection, MedRxiv. (2020) 2020.04.03.20052860. doi:10.1101/2020.04.03.20052860.

[25] OSF Preprints |Landscape Coronavirus Disease 2019 test (COVID-19 test) in vitro -- A comparison of PCR vs Immunoassay vs Crispr-Based test, (n.d.). https://osf.io/6eagn (accessed April 8, 2020).

[26] J.M. Rothberg, W. Hinz, T.M. Rearick, J. Schultz, W. Mileski, M. Davey, J.H. Leamon, K. Johnson, M.J. Milgrew, M. Edwards, J. Hoon, J.F. Simons, D. Marran, J.W. Myers, J.F. Davidson, A. Branting, J.R. Nobile, B.P. Puc, D. Light, T.A. Clark, M. Huber, J.T. Branciforte, I.B. Stoner, S.E. Cawley, M. Lyons, Y. Fu, N. Homer, M. Sedova, X. Miao, B. Reed, J. Sabina, E. Feierstein, M. Schorn, M. Alanjary, E. Dimalanta, D. Dressman, R. Kasinskas, T. Sokolsky, J.A. Fidanza, E. Namsaraev, K.J. McKernan, A. Williams, G.T. Roth, J. Bustillo, An integrated semiconductor device enabling non-optical genome sequencing, Nature. 475 (2011) 348–352. doi:10.1038/nature10242.

[27] D. Gosselin, M. Gougis, M. Baque, F.P. Navarro, M.N. Belgacem, D. Chaussy, A.G. Bourdat, P. Mailley, J. Berthier, Screen-Printed Polyaniline-Based Electrodes for the Real-Time Monitoring of Loop-Mediated Isothermal Amplification Reactions, Anal. Chem. 89 (2017) 10124–10128. doi:10.1021/acs.analchem.7b02394.

[28] D. Han, R. Chand, Y.S. Kim, Microscale loop-mediated isothermal amplification of viral DNA with real-time monitoring on solution-gated graphene FET microchip, Biosens. Bioelectron. 93 (2017) 220–225. doi:10.1016/j.bios.2016.08.115.

[29] C. Toumazou, L.M. Shepherd, S.C. Reed, G.I. Chen, A. Patel, D.M. Garner, C.J.A. Wang, C.P. Ou, K. Amin-Desai, P. Athanasiou, H. Bai, I.M.Q. Brizido, B. Caldwell, D. Coomber-Alford, P. Georgiou, K.S. Jordan, J.C. Joyce, M. La Mura, D. Morley, S. Sathyavruthan, S. Temelso, R.E. Thomas, L. Zhang, Simultaneous DNA amplification and detection using a pH-sensing semiconductor system, Nat. Methods. 10 (2013) 641–646. doi:10.1038/nmeth.2520.

[30] S. Xie, Y. Yuan, Y. Song, Y. Zhuo, T. Li, Y. Chai, R. Yuan, Using the ubiquitous pH meter combined with a loop mediated isothermal amplification method for facile and sensitive detection of Nosema bombycis genomic DNA PTP1, Chem. Commun. 50 (2014) 15932–15935. doi:10.1039/c4cc06449f.

[31] E. González-González, G. Trujillo-de Santiago, I.M. Lara-Mayorga, S.O. Martínez-Chapa, M.M. Alvarez, Portable and accurate diagnostics for COVID-19: Combined use of the miniPCR thermocycler and a well-plate reader for SARS-CoV-2 virus detection, PLoS One. 15 (2020) e0237418. doi:10.1371/journal.pone.0237418.

[32] Y.-P. Wong, S. Othman, Y.-L. Lau, S. Radu, H.-Y. Chee, Loop-mediated isothermal amplification (LAMP): a versatile technique for detection of micro-organisms, J. Appl. Microbiol. 124 (2018) 626–643. doi:10.1111/jam.13647.

[33] K. Nawattanapaiboon, E. Pasomsub, P. Prombun, A. Wongbunmak, A. Jenjitwanich, P. Mahasupachai, P. Vetcho, C. Chayrach, N. Manatjaroenlap, C. Samphaongern, T. Watthanachockchai, P. Leedorkmai, S. Manopwisedjaroen, R. Akkarawongsapat, A. Thitithanyanont, M. Phanchana, W. Panbangred, S. Chauvatcharin, T. Srikhirin, Colorimetric reverse transcription loop-mediated isothermal amplification (RT-LAMP) as a visual diagnostic platform for the detection of the emerging coronavirus SARS-CoV-2, Analyst. 146 (2021) 471–477. doi:10.1039/d0an01775b.

[34] M. Cohen, R. Khalaila, Saliva pH as a biomarker of exam stress and a predictor of exam performance, J. Psychosom. Res. 77 (2014) 420–425. doi:10.1016/j.jpsychores.2014.07.003.

[35] S. Baliga, S. Muglikar, R. Kale, Salivary pH: A diagnostic biomarker, J. Indian Soc. Periodontol. 17 (2013) 461–465. doi:10.4103/0972-124X.118317.

[36] E. Gonzalez-Gonzalez, I.M. Lara-Mayorga, A. Garcia-Rubio, C.E. Garciamendez-Mijares, G.E. Guerra-Alvarez, G. Garcia-Martinez, J.A. Aguayo-Hernandez, Y.-S. Zhang, S.O. Martinez-Chapa, G. Trujillo-de Santiago, M.M. Alvarez, Scaling diagnostics in times of COVID-19: Rapid prototyping of 3D-printed water circulators for Loop-mediated Isothermal Amplification (LAMP) and detection of SARS-CoV-2 virus, Cold Spring Harbor Laboratory Press, 2020. doi:10.1101/2020.04.09.20058651.

[37] G. Papadakis, A. Pantazis, N. Fikas, S. Chatziioannidou, K. Michaelidou, V. Pogka, M. Megariti, M. Vardaki, K. Giarentis, J. Heaney, E. Nastouli, T. Karamitros, A. Mentis, S. Agelaki, E. Gizeli, Real-time colorimetric LAMP methodology for quantitative nucleic acids detection at the point-of-care, BioRxiv. (2020) 2020.07.22.215251. doi:10.1101/2020.07.22.215251.

[38] M. Janíková, J. Hodosy, P. Boor, B. Klempa, P. Celec, Loop□mediated isothermal amplification for the detection of SARS□CoV□2 in saliva, Microb. Biotechnol. 14 (2021) 307–316. doi:10.1111/1751-7915.13737.

