## supplementary material for "Portable and label-free quantitative Loop-mediated Isothermal Amplification (qLAMP) for reliable COVID-19 diagnostics in 3 minutes: An Arduino-based detection system assisted by a pH microelectrode"

**Supplemental information**

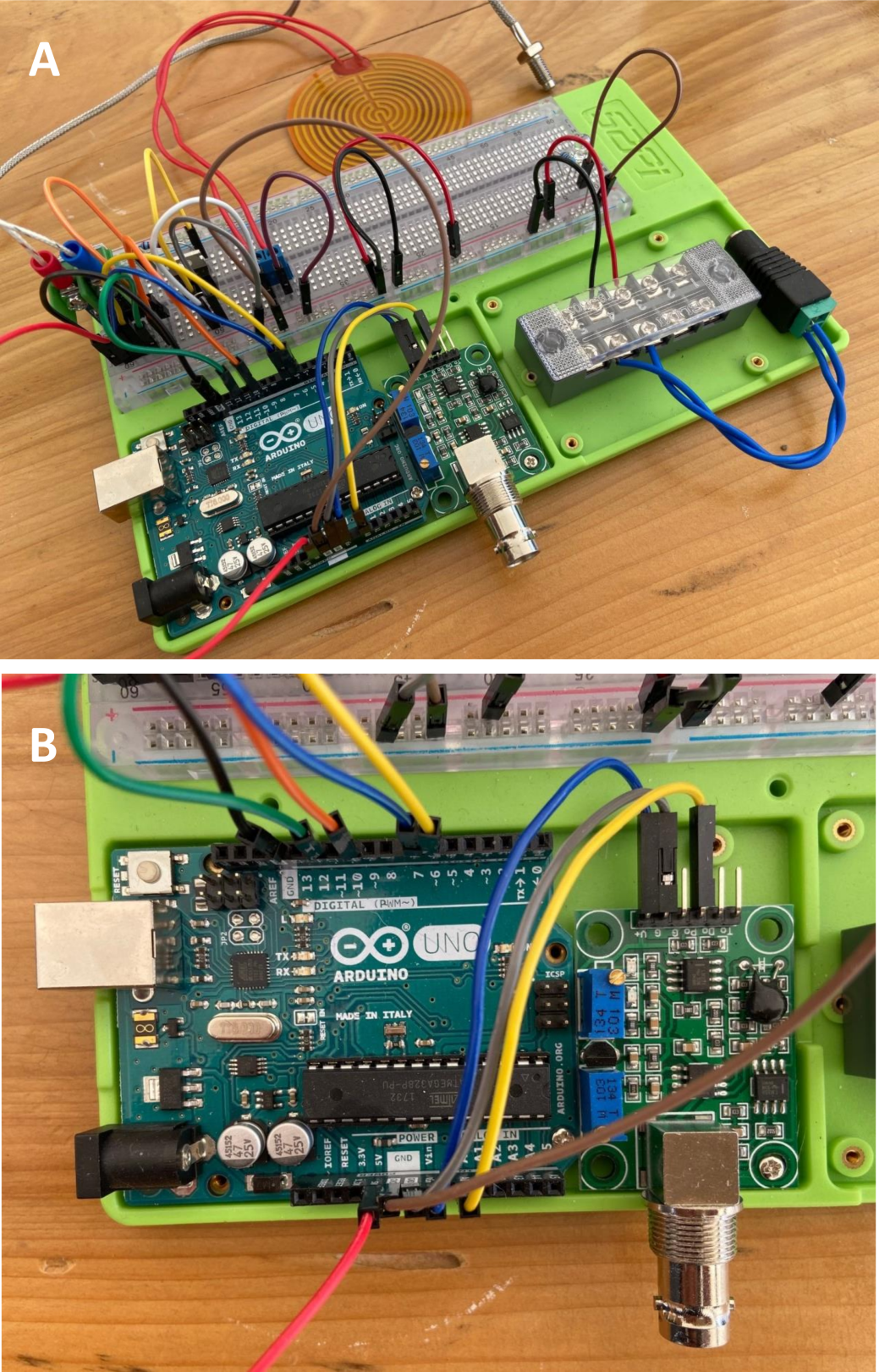

**S1_Figure.** Actual image of the Arduino-based proportional integral derivative (PID) temperature controller and the online pH/electric potential monitoring system used in our experiments.

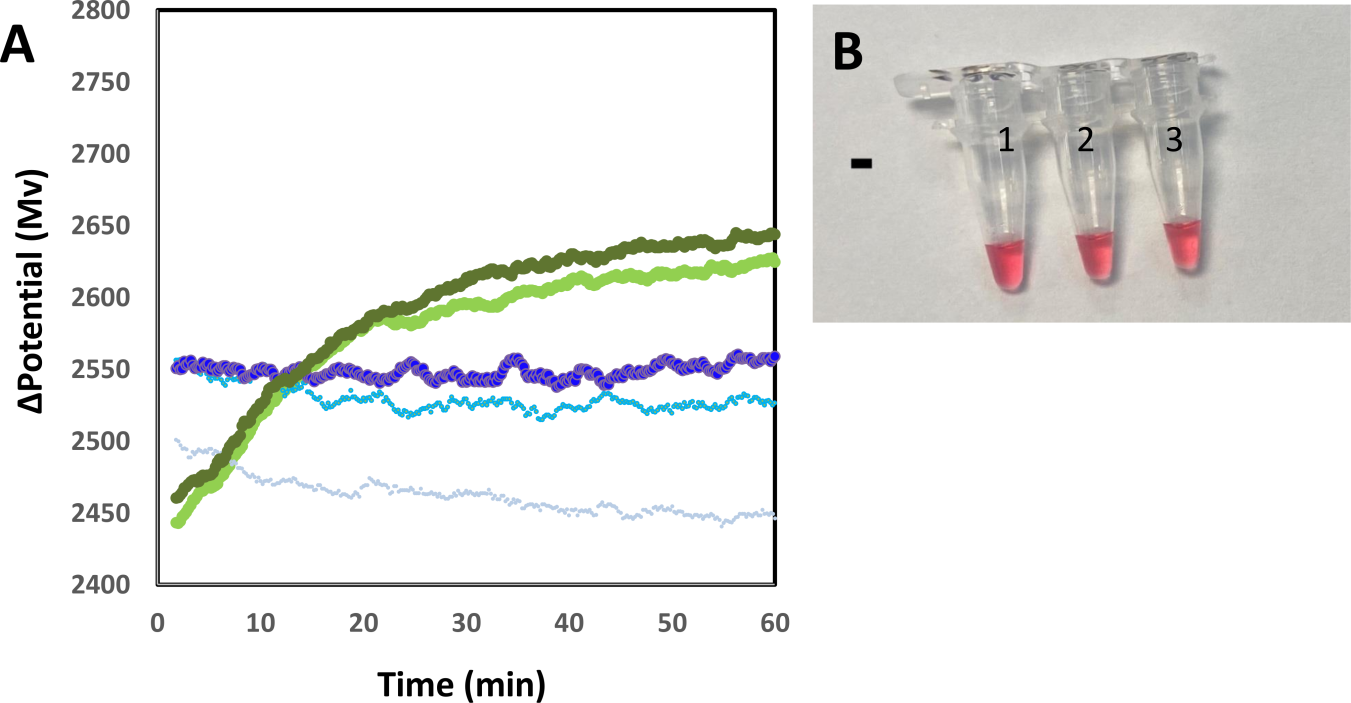

**S2_Figure.** Readings associated with negatives samples. (A) Readings of electric potential versus time are associated with three amplification-negative samples (samples that contained Loop-mediated Isothermal Amplification (LAMP) reaction mix but no primers or SARS-CoV-2 genetic material; green curves) and two negative samples (samples that contained reaction mix and primers but no SARS-CoV-2 genetic material; blue lines). (B) Actual images from three negative amplification samples (i.e., containing no primers).

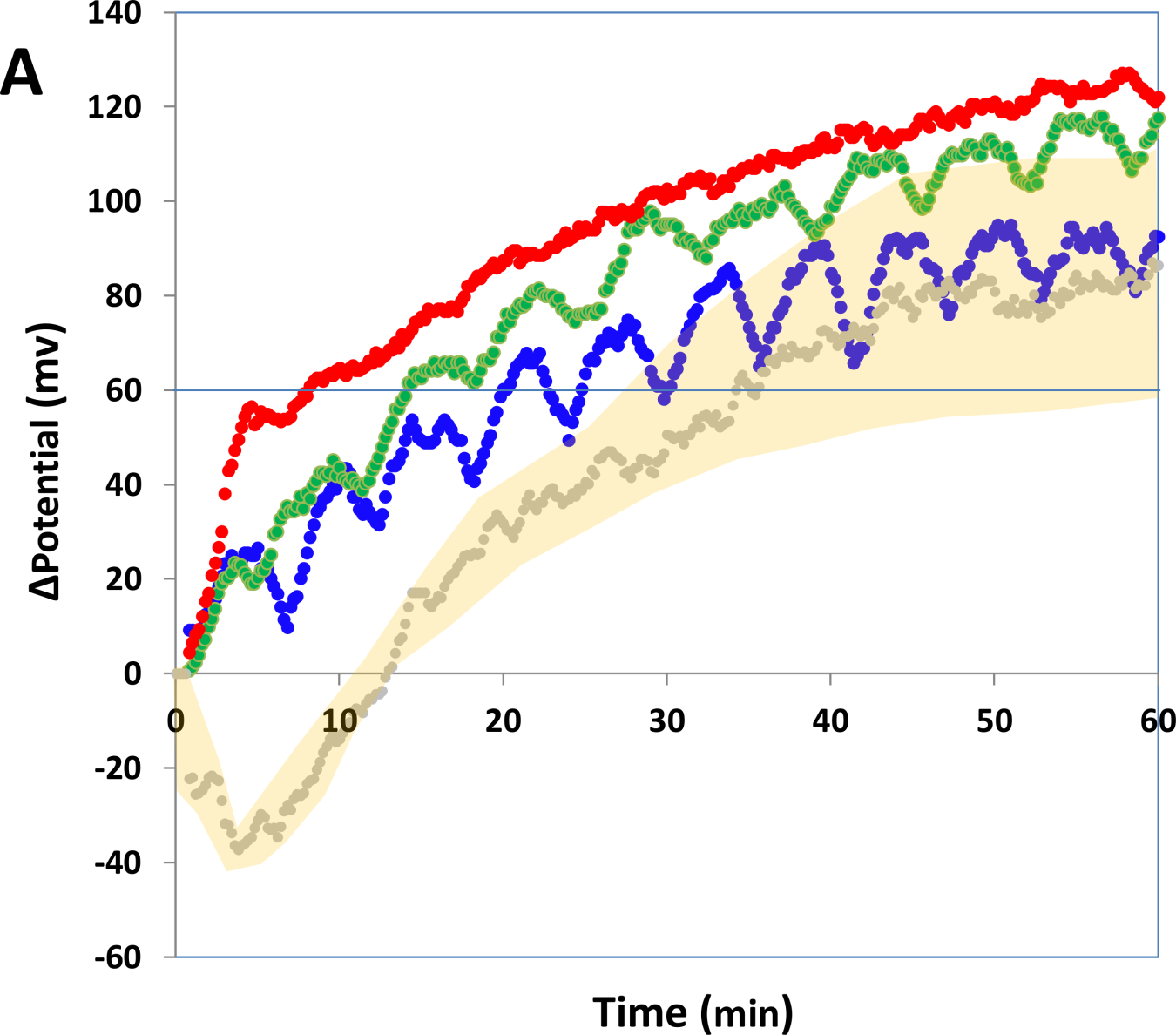

**S3_Figure.** Progression of the electric potential with time for negative samples (gray curve) and positive samples containing 100 (blue symbols), 1000 (green symbols), and 10,000 copies of synthetic SARS-CoV-2 genetic material (red symbols). Readings were collected at suboptimal sampling rates, which led to a deficient definition of the initial slope. The oscillation of the signal readings visible in two of the plots is related to oscillations in the temperature due to the use of an on-off temperature strategy for temperature control. The yellow shaded area indicates the dispersion associated with the readings of potential in negative samples in different independent experiments (n=3). All curves were normalized by subtracting the first ascending value of potential after placing the electrode in the reaction mix.

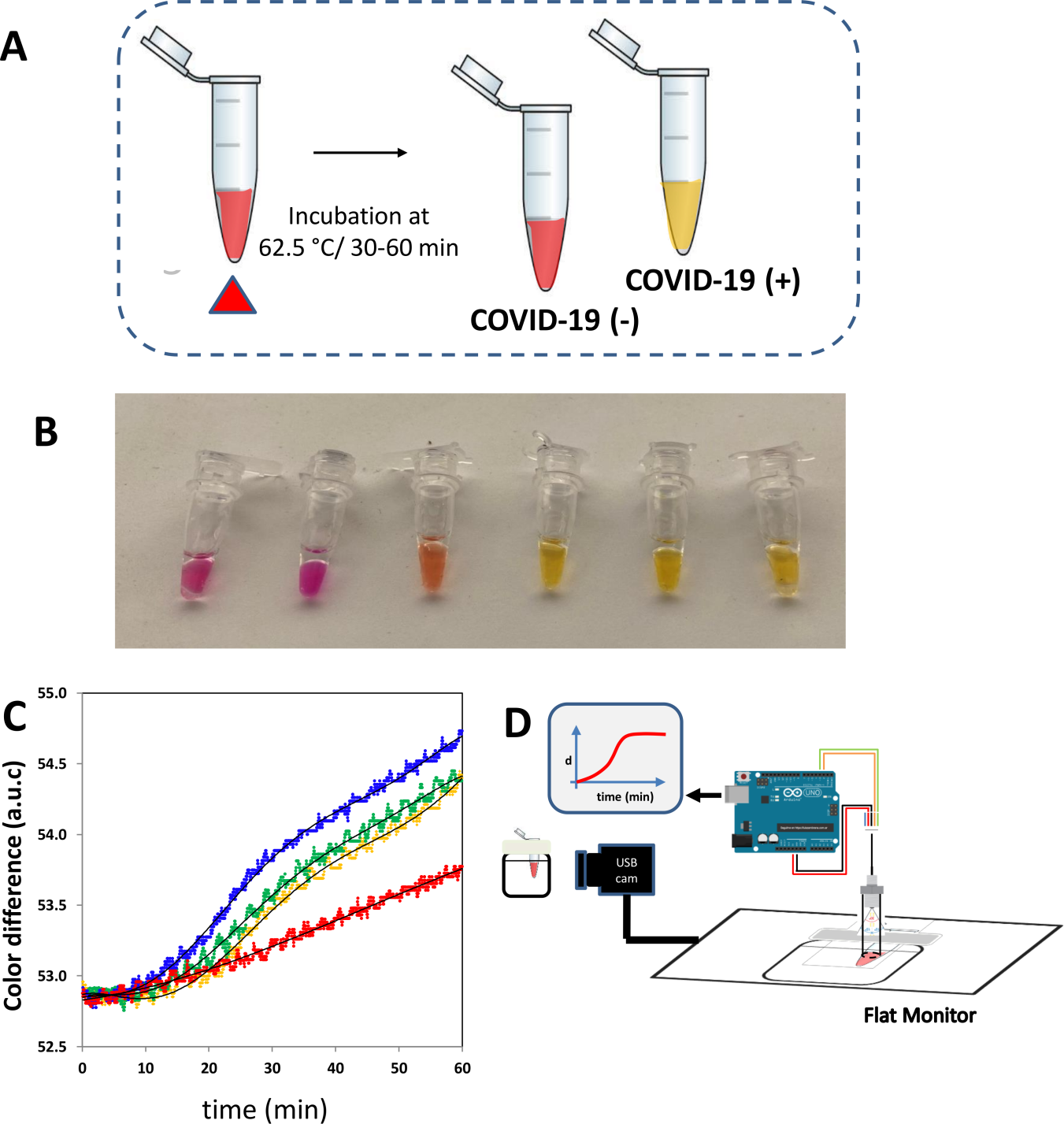

**S4_Figure. Real-time determination of color changes.** Colorimetric Loop-mediated Isothermal Amplification (LAMP) is commonly implemented as a final-point technique based on a change in color associated with a change in the pH of the reaction mix. The reaction mix is added to phenol red (threshold pH value ~ 6.8). In a weakly buffered solution, the pH changes during the amplification (specific or non-specific) and this change in pH induces a (B) progressive change in color that can be observed with the naked eye. (C) This evolution of the color can be monitored in real time under conditions of controlled illumination using commercial sensors (e.g., EZO sensor from Atlas Co., USA) and a USB camera. (D) Progression of the color difference in the sample between red (initial state) and yellow (final state) with added copies of SARS‑CoV‑2 genetic material: 0 (green curve; red), 100 (yellow curve), 1,000 (green curve), and 10,000 (blue curve).

**Table S1.** Average electric potential associated with buffers of different pH values. The standard deviation and the coefficients of variance of the potential determinations, as determined during online determinations of 60 minutes (n= 1800), are also reported.

| pH value | Pot (mV) | STD | CV (%) |
| --- | --- | --- | --- |
| 6.00 | 2891.32 | 2.76 | 0.10 |
| 6.40 | 2843.71 | 6.91 | 0.24 |
| 6.80 | 2777.02 | 1.38 | 0.05 |
| 7.20 | 2713.54 | 1.38 | 0.05 |
| 7.60 | 2639.10 | 2.76 | 0.10 |
| 8.00 | 2587.58 | 6.21 | 0.24 |

**Supplemental file S1: Arduino code**

**Arduino code**

(version1: considers the use of a of a Dallas-thermocouple)

// Arduino IDE code for the simultaneous control of temperature in a setpoint and determination of //electrical potential in the reaction mix during LAMP or PCR amplification.

// This version considers the use of a Dallas-thermocouple

// This program was assembled by Mario Moisés Alvarez at Tecnológico de Monterrey, México.

// To this aim, sections of code from various open sources were used:

//OPEN USE; PLEASE FOLLOW THESE RECOMMENDATIONS TO PROPERLY AKNOWLEDGE OUR LAB

// This code is intended for open use. If used, please cite the corresponding article if used.

// Please also acknowledge the Alvarez-Trujillo Lab, at Tecnológico de Monterrey, México.

// <http://www.alvareztrujillolab.com/>

// To run this code, several Arduino libraries must be downloaded and made accessible:

// --- <DallasTemperature.h> --- (for sinc with thermocouple) //<https://github.com/collin80/dallas_temperature_control/blob/master/DallasTemperature.h>

// --- <OneWire.h> --- (for communication) <https://github.com/PaulStoffregen/OneWire>

// --- <PID_v1.h> --- (for PID control) <https://github.com/br3ttb/Arduino-PID-Library>

//***************************************************************

// Arduino PID Library - Version 1.2.1

// by Brett Beauregard <> brettbeauregard.com

//

// This Library is licensed under the MIT License

//*************************************************************

// We acknowledge the contribution of the authors of these libraries that enable this code

#include <PID_v1.h>

#include <Arduino.h>

#include <OneWire.h>

#include <DallasTemperature.h>

OneWire ourWire(4); //Se establece el pin 4 como bus OneWire

DallasTemperature sensors(&ourWire); //Se declara una variable u objeto para nuestro sensor

const int adcPin = A0;

const int sampleRate = 1;

double Setpoint, Input, Output;

#define Kp 22.2

#define Ki 0.00

#define Kd 0.00

PID ControlPID(&Input, &Output, &Setpoint, Kp, Ki, Kd, DIRECT);

//Variables

float temperature_read = 0.0;

float time = 0;

float PID_error = 0;

float Setpointzero = 0;

float phVoltage1; float phVoltage2; float phVoltage3; float phVoltage4; float phVoltage5;

void setup() {

pinMode(10,OUTPUT);

ControlPID.SetMode(AUTOMATIC);

ControlPID.SetSampleTime(sampleRate);

//Initialize SetPoint

//Setpoint to 62.5 degrees

Setpoint = 62.5;

Setpointzero= Setpoint;

// The value “6.5” is a correction factor to artificially increase the set point and compensate for heat

//losses in the incubator. Please modify accordingly to achieve your aimed set point.

Setpoint= Setpoint + 6.5;

Input=0;

Output=0;

phVoltage1 = 0.0;

phVoltage2 = 0.0;

phVoltage3 = 0.0;

phVoltage4 = 0.0;

phVoltage5 = 0.0;

Serial.begin(9600);

}

void loop() {

float average;

int adcValue = analogRead(adcPin);

float phVoltage = (float)adcValue * 5000 / 1024 ;

phVoltage5= phVoltage4;

phVoltage4= phVoltage3;

phVoltage3= phVoltage2;

phVoltage2= phVoltage1;

phVoltage1= phVoltage;

phVoltage= (phVoltage1+phVoltage2+phVoltage3+phVoltage4+phVoltage5)/5;

uint8_t i;

average = 0.0;

int val=0;

val = round(Output);

analogWrite(10,val);

sensors.requestTemperatures(); //Se envía el comando para leer la temperatura

float temp= sensors.getTempCByIndex(0); //Se obtiene la temperatura en ºC

// +1.5 is a correction factor that should be modified as needed to conciliate Tread with Treactor

temperature_read = temp +2.0;

// The value “2.0” is a correction factor that corrects for the difference in the temperature reading of //the thermocouple in the bulk of the liquid in the incubator and the actual temperature of the reaction //mix in the Eppendorf tube.

//Next we calculate the error between the setpoint and the real value

PID_error = Setpointzero - temperature_read;

Input= temperature_read;

ControlPID.Compute();

Serial.print("T :");

Serial.print(temperature_read,1);

Serial.print(" C ; Po = "); Serial.println(phVoltage, 3);

delay (942);

}

**Arduino code**

(version2: considers the use of a MAX6675 module for T sensing)

// Arduino IDE code for the simultaneous control of temperature in a setpoint and determination of //electrical potential in the reaction mix during LAMP or PCR amplification.

// This version of code considers the use of a MAX6675 module for T sensing

// This program was assembled by Mario Moisés Alvarez at Tecnológico de Monterrey, México.

// To this aim, sections of code from various open sources were used:

//OPEN USE; PLEASE FOLLOW THESE RECOMMENDATIONS TO PROPERLY AKNOWLEDGE OUR LAB

// This code is intended for open use. If used, please cite the corresponding article if used.

// Please also acknowledge the Alvarez-Trujillo Lab, at Tecnológico de Monterrey, México.

// <http://www.alvareztrujillolab.com/>

// To run this code, several Arduino libraries must be downloaded and made accessible:

// --- <SPI.h> --- (for sinc with thermocouple) <https://www.arduino.cc/en/reference/SPI>

// --- <Wire.h> --- (for communication) <https://www.arduino.cc/en/reference/wire>

// --- <PID_v1.h> --- (for PID control) <https://github.com/br3ttb/Arduino-PID-Library>

//***************************************************************

// Arduino PID Library - Version 1.2.1

// by Brett Beauregard <> brettbeauregard.com

//

// This Library is licensed under the MIT License

//*************************************************************

// We acknowledge to the authors of these libraries that enable this code

//	This embodiment of the program uses a MAX6675 module for temperature reading

// Max6675 Module ==>	Arduino

//	CS	==>	D6

//	SO	==>	D12

//	SCK	==>	D7

//	Vcc	==>	Vcc (5v)

//	Gnd	==>	Gnd

#include <Wire.h>

#include <PID_v1.h>

#include <Arduino.h>

const int adcPin = A0;

const int sampleRate = 1;

double Setpoint, Input, Output;

#define Kp 22.2

#define Ki 0.00

#define Kd 0.00

PID ControlPID(&Input, &Output, &Setpoint, Kp, Ki, Kd, DIRECT);

#include <SPI.h>

//We define the SPI pìns

#define MAX6675_CS	6

#define MAX6675_SO	12

#define MAX6675_SCK 7

#define NUMSAMPLES 5

//Variables

float temperature_read = 0.0;

float time = 0;

float PID_error = 0;

unsigned long now;

float Setpointzero = 0;

float phVoltage1; float phVoltage2; float phVoltage3; float phVoltage4; float phVoltage5;

int counter1 = 0;

void setup() {

pinMode(10,OUTPUT);

ControlPID.SetMode(AUTOMATIC);

ControlPID.SetSampleTime(sampleRate);

//Initialize SetPoint

//Setpoint to 62.5 degrees

Setpoint = 62.5;

Setpointzero= Setpoint;

// The value “6.5” is a correction factor to artificially increase the set point and compensate for heat

//losses in the incubator. Please modify accordingly to achieve your aimed set point.

Setpoint= Setpoint + 6.5;

Input=0;

Output=0;

phVoltage1 = 0.0;

phVoltage2 = 0.0;

phVoltage3 = 0.0;

phVoltage4 = 0.0;

phVoltage5 = 0.0;

Serial.begin(9600);

}

void loop() {

float average;

int adcValue = analogRead(adcPin);

float phVoltage = (float)adcValue * 5000 / 1024 ;

phVoltage5= phVoltage4;

phVoltage4= phVoltage3;

phVoltage3= phVoltage2;

phVoltage2= phVoltage1;

phVoltage1= phVoltage;

phVoltage= (phVoltage1+phVoltage2+phVoltage3+phVoltage4+phVoltage5)/5;

// Serial.println (phVoltage5);

// Serial.println (phVoltage4);

// Serial.println (phVoltage3);

// Serial.println (phVoltage2);

// Serial.println (phVoltage1);

uint8_t i;

average = 0.0;

int val=0;

// First we read the real value of temperature

// take N samples in a row, with a slight delay

for (i=0; i< NUMSAMPLES; i++) {

samples[i] = readThermocouple();

//	Serial.println(readThermocouple());

delay(10);

average /= NUMSAMPLES; // calculate average

}

val = round(Output);

analogWrite(10,val);

temperature_read = readThermocouple()+2.0;

// The value “2.0” is a correction factor that corrects for the difference in the temperature reading of //the thermocouple in the bulk of the liquid in the incubator and the actual temperature of the reaction //mix in the Eppendorf tube.

//Next we calculate the error between the setpoint and the real value

PID_error = Setpointzero - temperature_read;

//Serial.println(PID_error,1);

Input= temperature_read;

ControlPID.Compute();

// Serial.println(val);

Serial.print("T :");

Serial.print(temperature_read,1);

Serial.print(" C ; Po = "); Serial.println(phVoltage, 3);

counter1 = counter1 + 1;

if (counter1 == 5) mV_baseline = phVoltage;

if (counter1 > 5) {

deltamV = phVoltage - mV_baseline;

}

delay (942);

}

double readThermocouple() {

uint16_t v;

pinMode(MAX6675_CS, OUTPUT);

pinMode(MAX6675_SO, INPUT);

pinMode(MAX6675_SCK, OUTPUT);

digitalWrite(MAX6675_CS, LOW);

delay(1);

// Read in 16 bits,

// 15	= 0 always

// 14..2 = 0.25 degree counts MSB First

// 2	= 1 if thermocouple is open circuit

// 1..0 = uninteresting status

v = shiftIn(MAX6675_SO, MAX6675_SCK, MSBFIRST);

v <<= 8;

v |= shiftIn(MAX6675_SO, MAX6675_SCK, MSBFIRST);

digitalWrite(MAX6675_CS, HIGH);

if (v & 0x4)

{

// Bit 2 indicates if the thermocouple is disconnected

return NAN;

}

// The lower three bits (0,1,2) are discarded status bits

v >>= 3;

// The remaining bits are the number of 0.25 degree (C) counts

return v*0.25;

}
